# Prediction of type 2 diabetes mellitus onset using logistic regression-based scoreboards

**DOI:** 10.1101/2020.08.02.20165092

**Authors:** Yochai Edlitz, Eran Segal

## Abstract

Type 2 diabetes mellitus (T2DM) accounts for ∼90% of all cases of diabetes which are estimated with an annual world death rate of 1.6 million in 2016. Early detection of T2D high-risk patients can reduce the incidence of the disease through a change in lifestyle, diet, or medication. Since populations of lower socio-demographic status are more susceptible to T2D and might have limited resources for laboratory testing, there is a need for accurate yet accessible prediction models based on non-laboratory parameters. This paper introduces one non-laboratory model which is highly accessible to the general population and one highly precise yet simple laboratory model. Both models are provided as an accessible scoreboard form and also as a logistic regression model. We based the models on data from 44,879 non-diabetic, UK Biobank participants, aged 40-65, predicting the risk of T2D onset within the next 7.3 years (SD 2.3). The non-laboratory prediction model for T2DM onset probability incorporated the following covariates: sex, age, weight, height, waist, hips-circumferences, waist-to-hip Ratio (WHR) and Body-Mass Index (BMI). This logistic regression model achieved an Area Under the Receiver Operating Curve (auROC) of 0.82 (0.79-0.85 95% CI) and an odds ratio (OR) between the upper and lower prevalence deciles of x77 (28-98). We further analysed the contribution of laboratory-based parameters and devised a blood-test model based on just five blood tests. In this model, we included age, sex, Glycated Hemoglobin (HbA1c%), reticulocyte count, Gamma Glutamyl-Transferase, Triglycerides, and HDL cholesterol to predict T2D onset. This logistic-regression model achieved an auROC of 0.89 (0.86-0.91) and a deciles’ OR of x87 (27-152). Using the scoreboard results, the Anthropometrics model classified three risk groups, a group with 1%(1-2%); a group with 9% (7-11%) probability, and a group with a 15% (7-23%) risk of developing T2D. The Five blood tests scoreboard model, further classified into four risk groups: 0.9% (0.7%-1%); 8%(6-11%); 18%(14-22%) and a high risk group of 38%(23-54%) of developing T2D. We analysed several more comprehensive models which included genotyping data and other environmental factors and found that it did not provide cost efficient benefits over the five blood tests model. The Five blood tests and anthropometric models, both in their logistic regression form and scoreboard form, outperform the commonly used non-laboratory models, the Finnish Diabetes Risk Score (FINDRISC) and the German Diabetes Risk Score (GDRS). When trained using our data, the FINDRISC achieved an auROC of 0.75 (0.71-0.78), and the GDRS auROC resulted in 0.58 (0.54-0.62), respectively.

## 1. Introduction

Diabetes mellitus is defined as a group of diseases characterised by symptoms of chronic hyperglycemia. It is becoming one of the world’s most challenging epidemics. The prevalence of T2D has increased from 4.7% in 1980 to 8.5% in 2014. An estimated 1.6 million deaths were directly caused by diabetes in 2016. T2D is generally characterised by insulin resistance, resulting in hyperglycemia, and it accounts for ∼90% of all diabetes cases ^1,2^.

In recent years, the prevalence of diabetes has been rising more rapidly in low and middle-income countries (LMICs) than in high-income countries^3^. In 2014 Beagley et al. estimated that 45.8% or 174.8 million of all diabetes cases in adults are undiagnosed. 83.8% of all cases of undiagnosed diabetes are in LMICs ^4^, where laboratory diagnosis testing is limited for some of the populations in these countries^5^.

According to several studies, a healthy diet, regular physical activity, maintaining normal body weight and avoiding tobacco use can prevent or delay T2D onset ^3,6,7,8,9^. A screening tool that can identify individuals at risk will enable a lifestyle or medication intervention. Ideally, such a screening tool should be accurate, simple and low cost. It should also be easily available, allowing populations who have difficulty accessing laboratories to be screened by other means.

Several such tools are in use today ^10,11,12^. The Finnish Diabetes Risk Score (FINDRISC), a commonly used, non-invasive T2D risk-score model, estimates patients aged between 35 and 64 developing T2D within the next ten years. FINDRISC was created based on a prospective cohort of 4,746 and 4,615 individuals in Finland in 1987 and 1992. The FINDRISC model uses gender, age, Body Mass Index (BMI), blood pressure medications, a history of high blood glucose, physical activity, daily consumption of fruits, berries, or vegetables and family history of Diabetes as the parameters for the model. The FINDRISC might be used as a scoreboard model or a logistic regression (LR) model ^13,14,15^.

Another commonly used prediction model is the German Diabetes Risk Score (GDRS), which estimates a five-year risk for developing T2D. The GDRS is based on 9,729 men and 15,438 women aged 35-65 years from the European Prospective Investigation into Cancer and Nutrition (EPIC)-Potsdam study ^16^. The GDRS is a Cox regression model based on age, height, waist circumference, the prevalence of hypertension (yes/no), smoking behaviour, physical activity, moderate alcohol consumption, coffee consumption, intake of whole-grain bread, intake of red meat, and parent and sibling history of T2D ^17,18^. This model, too, can be assigned as an accessible scoreboard model.

The objective of the present research was to develop clinically usable models which are easy to use and highly predictive of T2D onset. We developed two simple models and compared their predictive power to the highly esteemed FINDRISC and GDRS as our baseline. We trained all models on a training data set and tested them on the holdout data set, taken from the UK Biobank (UKB) observational study cohort. We based one of the models on easily accessible anthropometric measures and the other on an invasive laboratory test using only five blood samples.As our models were trained and evaluated using the UKB database, they are most relevant for the U.K. population aged 40-65. Still, they can also be used for people similar to our research cohort (as presented in Table 1) and might be adapted to additional populations. Both models are given both in their logistic regression form and as accessible scoreboards.

**Table 1.**
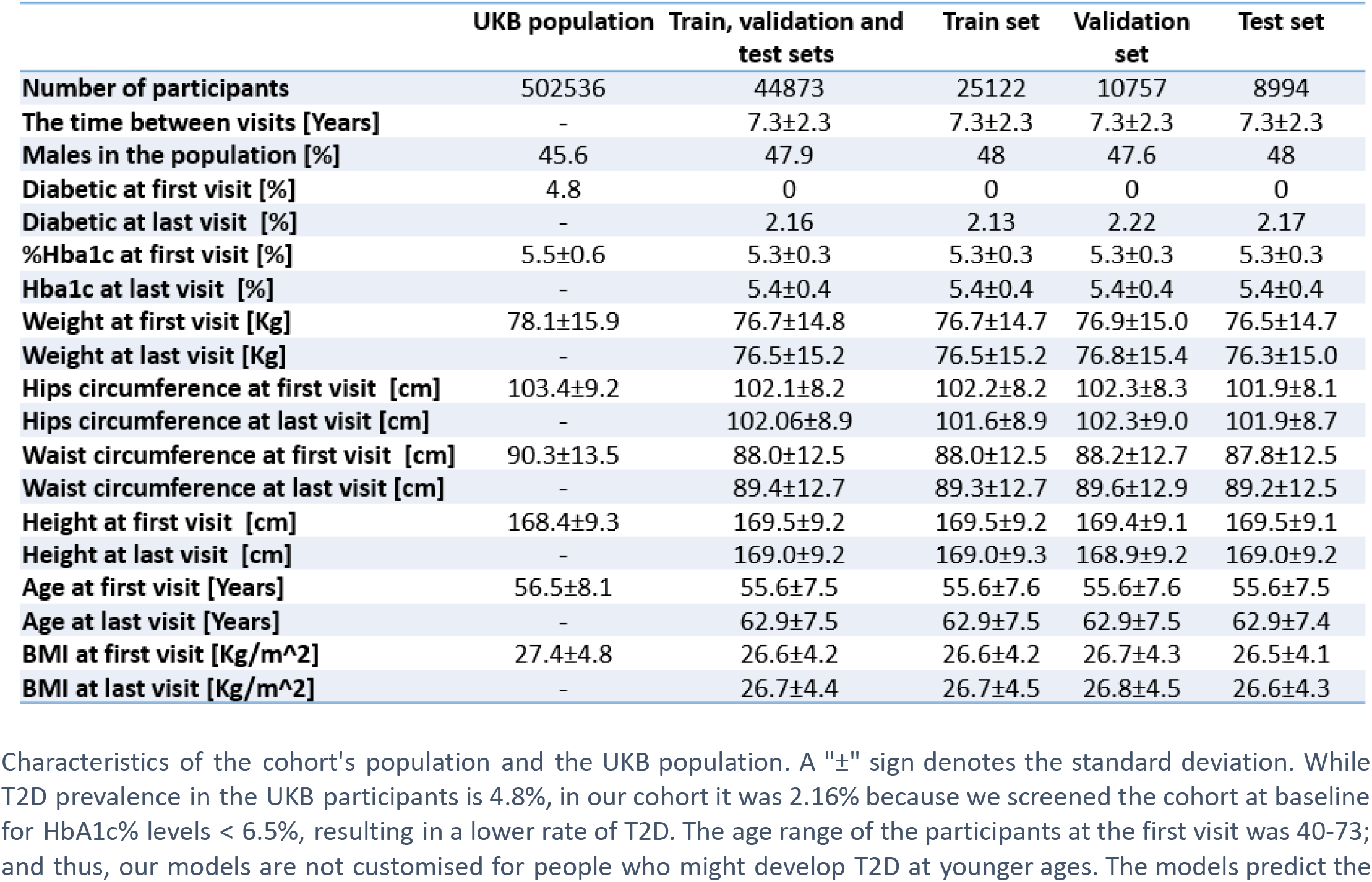
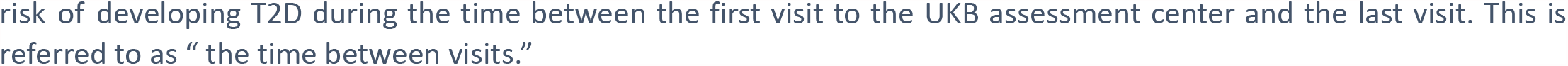
Cohort statistical data.

|  | UKB population | Train, validation and test sets | Train set | Validation set | Test set |
| --- | --- | --- | --- | --- | --- |
| **Number of participants** | 502536 | 44873 | 25122 | 10757 | 8994 |
| **The time between visits [Years]** | - | 7.3±2.3 | 7.3±2.3 | 7.3±2.3 | 7.3±2.3 |
| **Males in the population [%]** | 45.6 | 47.9 | 48 | 47.6 | 48 |
| **Diabetic at first visit [%]** | 4.8 | 0 | 0 | 0 | 0 |
| **Diabetic at last visit [%]** | - | 2.16 | 2.13 | 2.22 | 2.17 |
| **%Hba1c at first visit [%]** | 5.5±0.6 | 5.3±0.3 | 5.3±0.3 | 5.3±0.3 | 5.3±0.3 |
| **Hba1c at last visit [%]** | - | 5.4±0.4 | 5.4±0.4 | 5.4±0.4 | 5.4±0.4 |
| **Weight at first visit [Kg]** | 78.1±15.9 | 76.7±14.8 | 76.7±14.7 | 76.9±15.0 | 76.5±14.7 |
| **Weight at last visit [Kg]** | - | 76.5±15.2 | 76.5±15.2 | 76.8±15.4 | 76.3±15.0 |
| **Hips circumference at first visit [cm]** | 103.4±9.2 | 102.1±8.2 | 102.2±8.2 | 102.3±8.3 | 101.9±8.1 |
| **Hips circumference at last visit [cm]** | - | 102.06±8.9 | 101.6±8.9 | 102.3±9.0 | 101.9±8.7 |
| **Waist circumference at first visit [cm]** | 90.3±13.5 | 88.0±12.5 | 88.0±12.5 | 88.2±12.7 | 87.8±12.5 |
| **Waist circumference at last visit [cm]** | - | 89.4±12.7 | 89.3±12.7 | 89.6±12.9 | 89.2±12.5 |
| **Height at first visit [cm]** | 168.4±9.3 | 169.5±9.2 | 169.5±9.2 | 169.4±9.1 | 169.5±9.1 |
| **Height at last visit [cm]** | - | 169.0±9.2 | 169.0±9.3 | 168.9±9.2 | 169.0±9.2 |
| **Age at first visit [Years]** | 56.5±8.1 | 55.6±7.5 | 55.6±7.6 | 55.6±7.6 | 55.6±7.5 |
| **Age at last visit [Years]** | - | 62.9±7.5 | 62.9±7.5 | 62.9±7.5 | 62.9±7.4 |
| **BMI at first visit [Kg/m^2]** | 27.4±4.8 | 26.6±4.2 | 26.6±4.2 | 26.7±4.3 | 26.5±4.1 |
| **BMI at last visit [Kg/m^2]** | - | 26.7±4.4 | 26.7±4.5 | 26.8±4.5 | 26.6±4.3 |

Characteristics of the cohort's population and the UKB population. A "±" sign denotes the standard deviation. While T2D prevalence in the UKB participants is 4.8%, in our cohort it was 2.16% because we screened the cohort at baseline for HbA1c% levels < 6.5%, resulting in a lower rate of T2D. The age range of the participants at the first visit was 40-73; and thus, our models are not customised for people who might develop T2D at younger ages. The models predict the
risk of developing T2D during the time between the first visit to the UKB assessment center and the last visit. This is referred to as “the time between visits.”

## 2. Results

We analysed the data of 20,346 participants from the UK Biobank’s (UKB) cohort who revisited the UKB assessment centers during 2012-2013 and 48,705 participants who revisited the centers from 2014 onwards (see Figure 1 and Methods). During the screening process of our cohort, we kept the data of the participants who returned for a second or third visit, tested negative for T2D and were not treated for T2D. The final cohort sample included data of 44,873 participants, of whom 2.16% developed T2D during a follow-up period of 7.3±2.3 years (see Table 1, Figure 1A and Methods).

**Figure 1.**
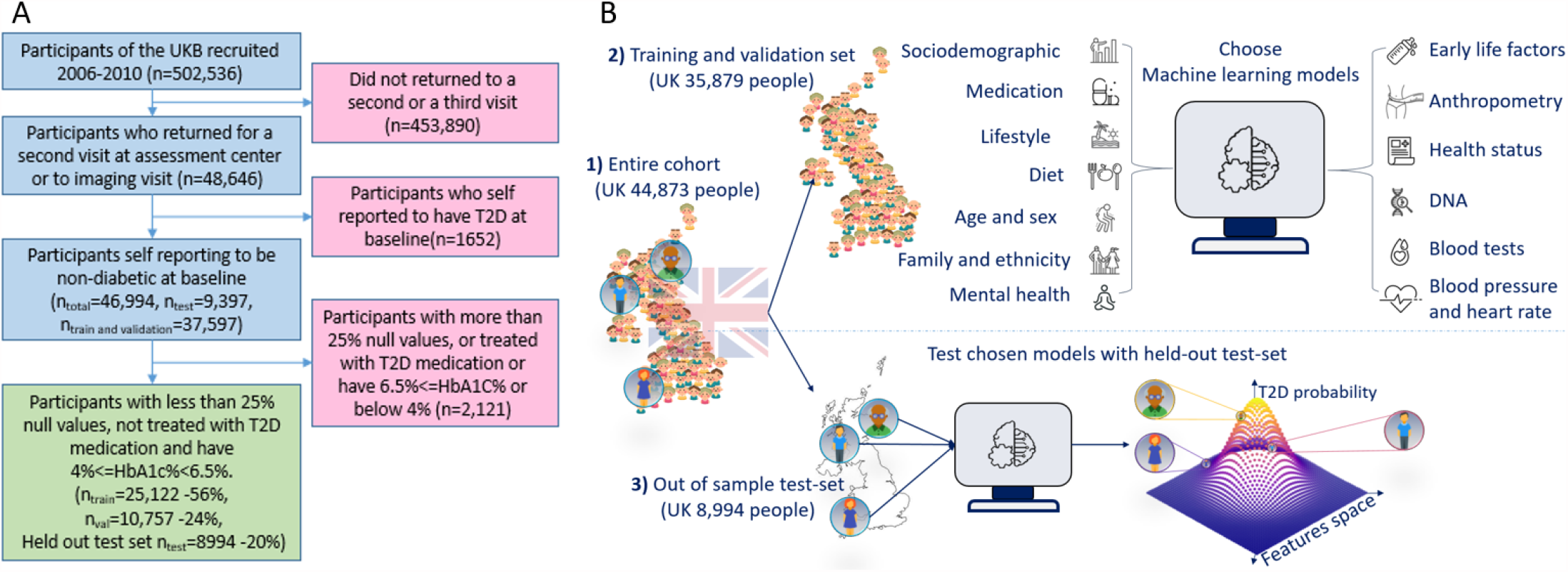
A flow chart of the cohort selection process and an illustrative figure of the model’s extraction. **A**. A flowchart which demonstrates the selection process of participants in this study. Participants who came for a repeated second or third visit were selected from the 502,536 participants of the UKB. Next, we excluded 1,652 participants who self-reported having T2D. We then split the data into 80% of the training and validation set, and 20% holdout test set. We excluded an additional 2,115 participants due to (1) having 25% or more missing values from the full feature list, (2) having HbA1c levels above or equal to 6.5%, or (3) being treated with Metformin or Insulin. Finally, the training set included 25,122 participants (56% of the cohort), the validation set included 10,757 (24% of the cohort), and the test set included a total of 8,994 participants (20% of the cohort). **B**. Process flow during training and testing of the models. We first split the data and kept a holdout test set. We later explored several models using the training and validation data sets. In the final stage, we compared the selected models using the holdout test set and reported the results. The output of the models is calibrated to predict the probability of a participant to develop T2D.

Before training the models, we partitioned our data into training, validation, and holdout test sets to avoid overfitting. The training dataset consisted of 25,122 participants, and the validation dataset included 10,757 participants. We explored the training and validation datasets to select the optimal features for our models. We used the holdout test set, which included 8,994 participants, to report the final models’ results (see Figure 1S and Methods).

### 2.1 Anthropometric based model

To provide an accessible, simple, non-laboratory and non-invasive T2D prediction model, we built anthropometric based ascoreboard model where a patient can easily mark its result in each of the scoreboard questions, consisting of the following eight parameters: age, sex, weight, height, hip circumference, waist circumference, body mass index (BMI) and the waist-to-hip ratio (WHR) (Figure 2A). The patient then sums up its final score which relates to one of three risk groups first group score range [1-70] has a 1% [1-2% 95% CI] of developing T2D; Second group, score range 71-83 predicts a 9%[7-11% 95%CI] of developing T2D; Third group 84-92 15% [7-23% 95%CI] of developing T2D (Figure 2C).

**Figure 2.**
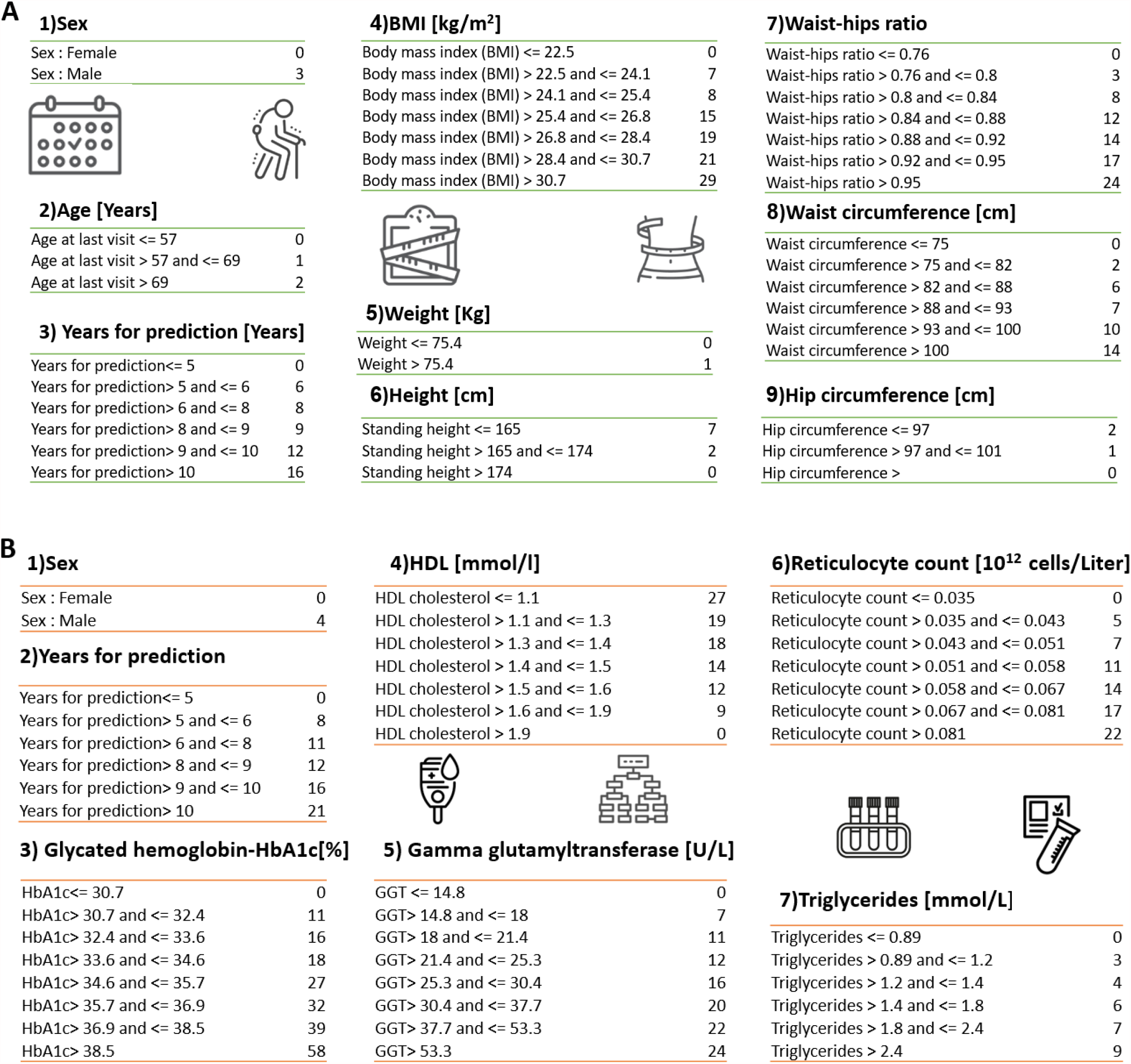
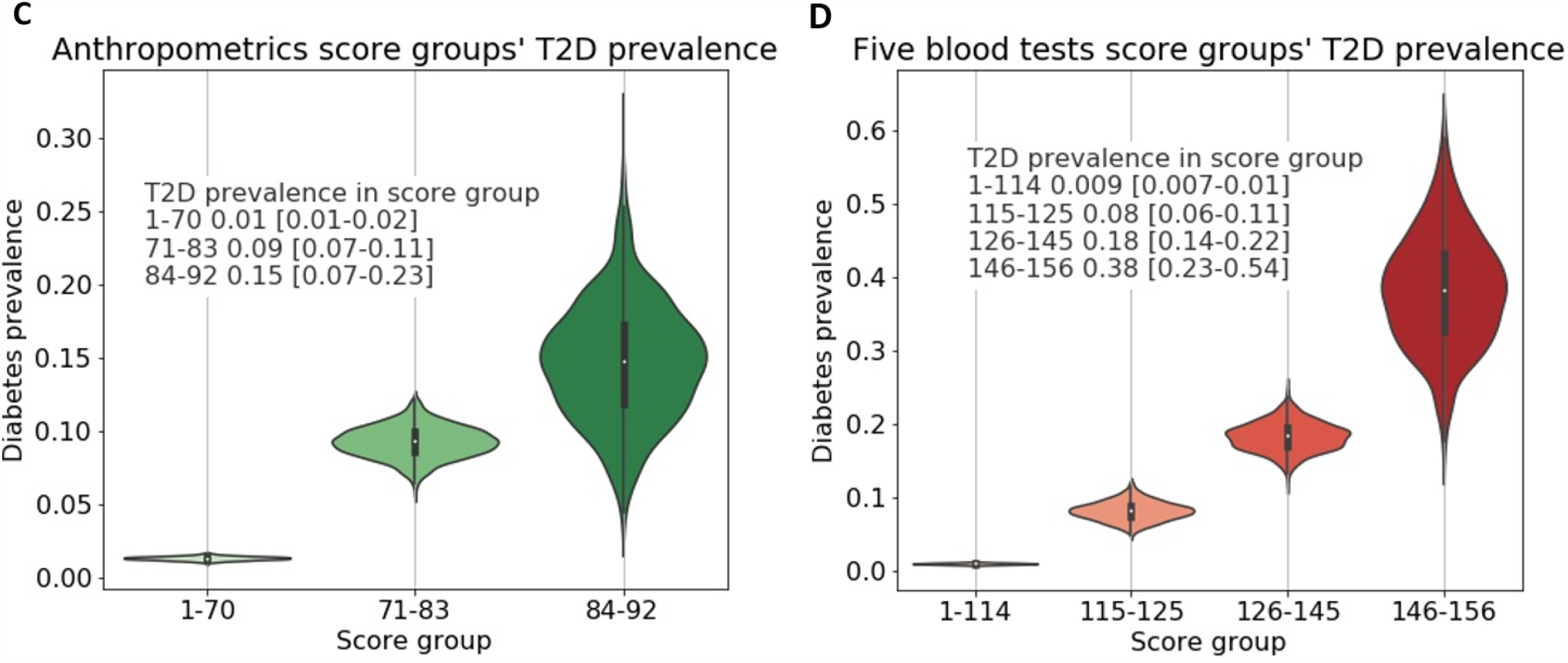
Anthropometrics and Blood tests scoreboards. A) Anthropometrics based scoreboard. Scoreboard, summing the scores of the various features provides a final score that is quantified into one of three risk groups. B) Five blood tests scoreboard. Summing the scores of the various features provides a final score that is quantified into one of four risk groups (Figure 2D). C) Anthropometrics scoreboards risk groups -first group score range [1-70] 1% [1-2%]95%CI of developing T2D; Second group, score range 71-83 predicts a 9% [7-11%] 95%CI of developing T2D.; Third group 84-92 15% [7-23%]95%CI of developing T2D. D) Five blood tests scoreboards risk groups -first group score range [1-114] <1% [0.7-1%]95%CI of developing T2D; Second group, score range 115-125 predicts an 8% [6-11%]95%CI of developing T2D.; Third group 126-145 18% [14-22%] 95%CI of developing T2D. Fourth group 146-156 predicts 38% [23-54%] 95%CI of developing T2D.

We also provide a similar model in its logistic regression form for more accurate computer aided results. Testing this model using the holdout test set, the logistic regression form of this model achieved an area under the receiver operating curve (auROC) of 0.82 (0.79-0.85) and an average precision score (APS) of 0.12 (0.09-0.16) at 95% CI). Using the model in its scoreboard form, we achieved an auROC of 0.81 (0.78-0.84) and an APS of 0.09 (0.07-0.12). Both model’s forms outperformed the two models which we used as a reference: the FINDRISC model, which has an auROC of 0.75 (0.71-0.78) and an APS of 0.07 (0.05-0.09), and the GDRS model, which has an auROC of 0.58 (0.54-0.62) and APS of 0.03 (0.02-0.04), see Figure 3A-B and Methods. With the cohort’s baseline prevalence of 2.17%, the LR anthropometric model achieved deciles’ OR of x77 (27.7-98.1), and its scoreboard form achieved deciles OR of x61 (17.7-101) compared to the FINDRISC x23 (6.80-70.4) and the x4.1 (1.75-9.24), see Figure 3C and Table 2. Analysing the models’ feature importance, the WHR and the BMI have the highest predictability in the anthropometric model due to their highest logs-odds-ratio (β) (Figure 3D). These two body habitus measures are commonly mentioned in the literature as indicators associated with chronic illness ^19,20,21,22^.

**Table 2.** Comparing models main results.

| Label | Model type | APS | AUROC | Deciles prevalence odds ratio |
| --- | --- | --- | --- | --- |
| **GDRS SA** | Cox regression | 0.03 (0.02-0.04) | 0.58 (0.54-0.62) | 4.1 (1.75-9.24) |
| **FINDRISC LR** | Scoreboard | 0.07 (0.05-0.09) | 0.75 (0.71-0.78) | 23 (6.80-70.4) |
| **Anthropometry** | Scoreboard | 0.09 (0.07-0.12) | 0.81 (0.78-0.84) | 61 (17.7-101) |
| **Anthropometry** | Logistic regression | 0.12 (0.09-0.16) | 0.82 (0.79-0.85) | 77 (27.7-98.1) |
| **Five blood tests** | Scoreboard | 0.18 (0.14-0.23) | 0.88 (0.85-0.90) | 78 (23.4-139) |
| **Five blood tests** | Logistic regression | 0.26 (0.20-0.33) | 0.89 (0.86-0.91) | 87 (26.7-152) |
| **Full blood tests** | Logistic regression | 0.32 (0.25-0.39) | 0.91 (0.89-0.93) | 117 (36.5-163) |
| **All features DT** | Gradient boosting decision trees | 0.34 (0.28-0.42) | 0.92 (0.90-0.94) | 133 (45.1-167) |

The values in parentheses indicate 95% CI. The deciles' OR is a measure of the ratio between T2D prevalence in the top risk score decile bin and the prevalence in the lowest decile bin. For the lowest decile, in case the actual prevalence in that bin was zero, we used a threshold of one diagnosed participant (see Methods).

**Figure 3.**
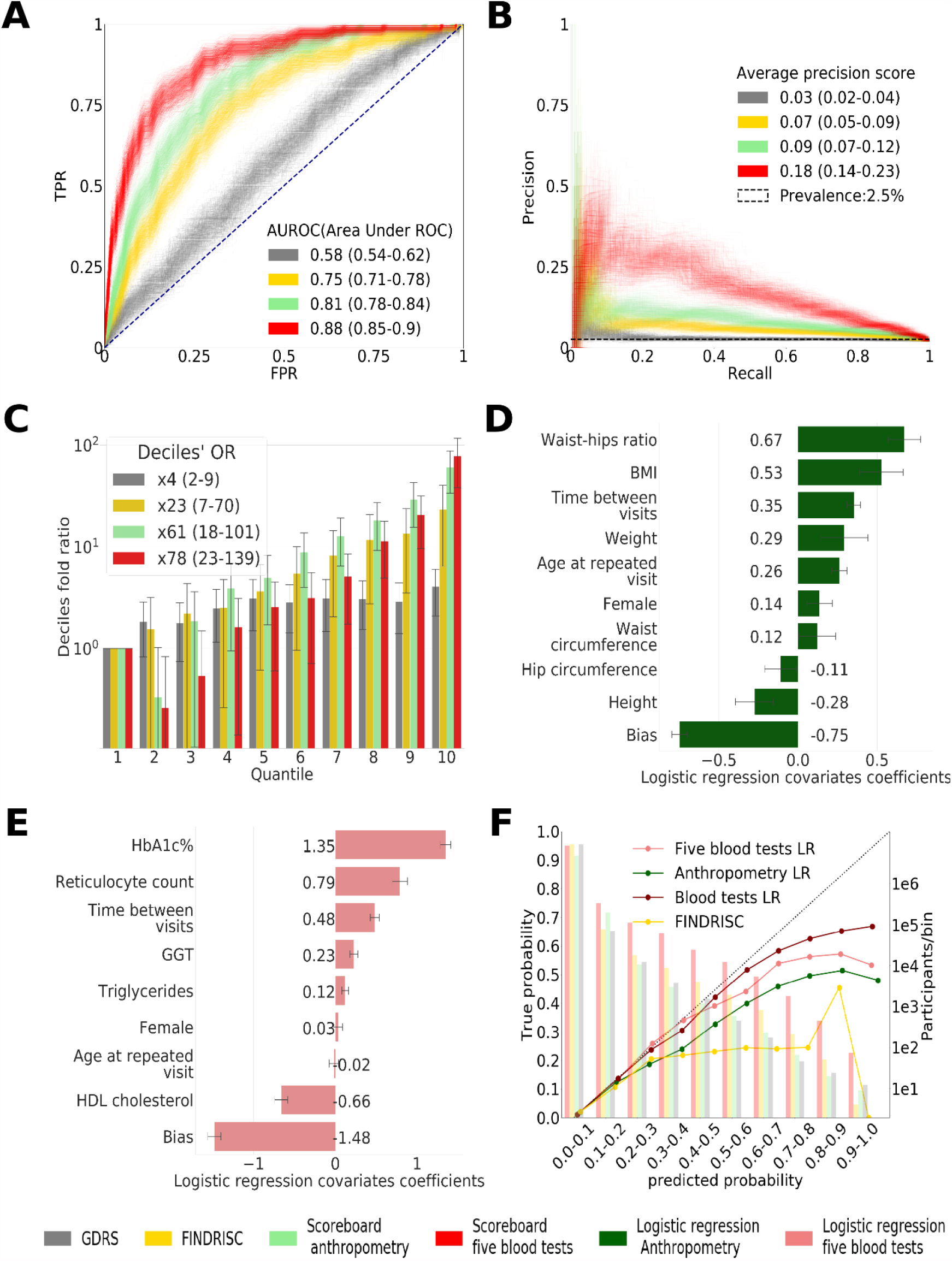
Main results calculated using 1000 bootstraps of the cohort population. Each point in the graphs represents a bootstrap iteration result. The color legend is shown at the bottom of the figure. **A**. ROC curves comparing the models developed in this research: a GBDT model of all features; logistic-regression models of five blood tests and the anthropometry based model compared to the well established GDRS and FINDRISC. **B**. Precision-Recall (P-R) curves, showing the precision versus the recall for each model, with the prevalence of the population marked with the dashed line. **C**. Deciles’ odds-ratio graph, the ratio of prevalence in each decile to the prevalence in the first decile. We bounded the prevalence in the first decile to be at least a tenth of the T2D prevalence in the full cohort. **D**. A feature importance graph of the logistic regression anthropometry model for a model with normalised features values. The bars indicate the standard deviation (SD) of the feature importance values. The top predictive features of this model are the body mass index (BMI) and waist to hip ratio (WHR). **E**. Feature importance graph of logistic regression Blood-tests model with SD bars. While the HbA1c% and Reticulocyte positively contribute to the T2D prediction, and HDL cholesterol lowers the T2D prediction probability, the information provided age and sex which is relevant for the prediction of T2D onset is overpowered by other features. **F**. A calibration plot of the anthropometry; five blood tests; full blood-test and the FINDRISC models. Calibration of the models’ predictions allow reporting the probability of developing T2D (see Methods). The calibration was performed using an isotonic regression method.

**Figure 4.**
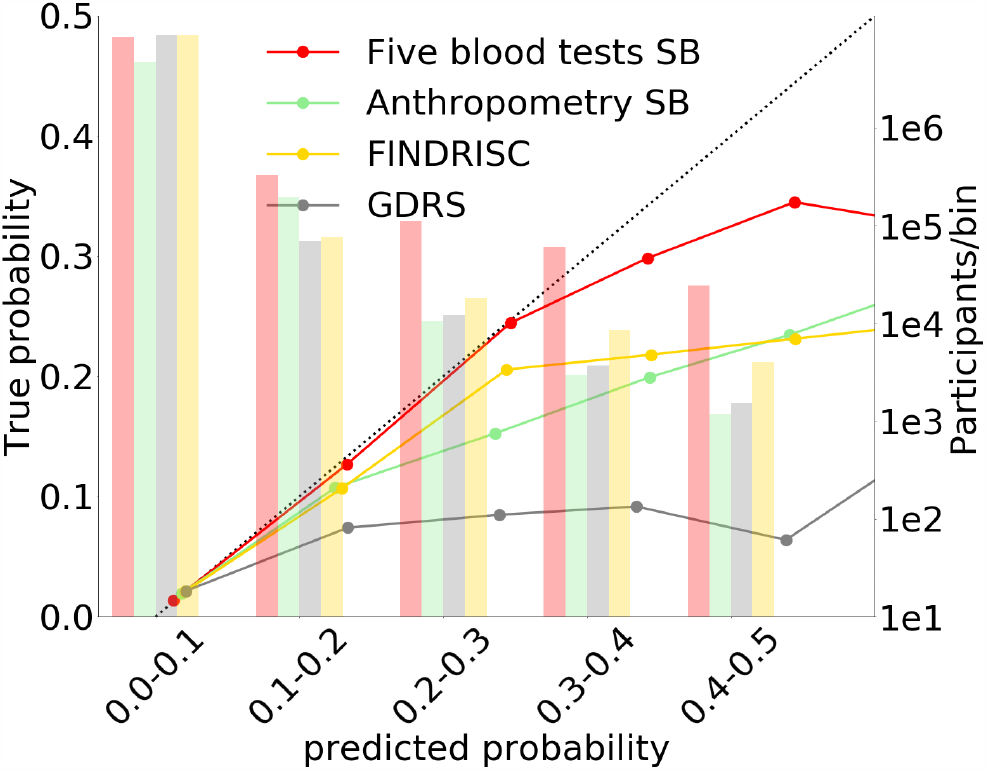
Anthropometric, five blood tests FINDRISC and GDRS scoreboards calibration graphs. While Five blood tests, Anbthropometrics and GDRS are monotonically rising, the FINDRISC model starts to decline after the third bin. The calibration of the Anthropometric and Five blood tests deteriorates compared to the continuous logistic regression model due to the scores quantisation effect.

### 2.2 five blood tests based model

In addition, for those cases where laboratory testing will be available, we developed a more accurate tool for predicting T2D onset. This tool uses five blood test scores as an input to a logistic regression model which we also simplified to a scoreboard model (Figure 2B,D). Using the Five blood tests scoreboard (Figure 2B), we bin the resulted scores into four groups: first group score range [1-114] has a 0.9% [0.7-1%] 95% CI probability of developing T2D; Second group, score range 115-125 predicts an 8% [6-11%] 95% CI probability of developing T2D; Third group score range 126-145 18% [14-22%] 95% CI of developing T2D; The fourth group score range is 146-156, participants in these score range has 35% [24-46%] 95% CI of developing T2D.

To derive this model, we started by a feature selection process from a full-feature GBDT model, using only the training and validation data sets. We clustered the features of this model into 13 categories such as lifestyle, diet, and anthropometrics. Based on this process, we concluded that the blood tests have higher predictability than the other clusters assessed. We thus trained a full blood test model using 59 blood tests that are available in the training dataset. Applying a recursive feature elimination process on the top 10 predictive features, we established the features of our final model, which is based on five blood tests (see Methods).

Using the five blood tests logistic regression model we achieved the following results for the test set: an auROC of 0.89 (0.86-0.91), an APS of 0.26 (0.2-0.33), and a deciles’ OR of x87 (27-152). When using the scoreboard model. we achieve an auROC of 0.88 (0.85-0.9), an APS of 0.18 (0.14-0.23), and a deciles’ OR of x78 (23-139) (Figure 3 A-C, Table 2). The five blood tests model results are superior to those of our non-laboratory anthropometric model, as well as those of the highly-esteemed FINDRISC and GDRS models (Figure 3 A-C, Table 2).

We then compared these results to those of a 59 blood tests input features of logistic regression model and to those of a GBDT model including 13 feature clusters, which consisted of 279 individual features available in the dataset and genetics data. These two models achieved an auROC of 0.91 (0.88-0.93) and 0.92 (0.9-0.94); an APS of 0.32 (0.25-0.39) and 0.34 (0.28-0.42); and a deciles’ OR of x117 (37-163) and x133(45-167), respectively.

The five blood tests that we used are the following: the Glycated Haemoglobin test(HbA1c%), which measures the average blood sugar for the past two to three months and which is one of the means to diagnose Diabetes; the Reticulocyte Count; the Gamma-Glutamyl Transferase Test (GGT); the HDL Cholesterol Test, and the Triglycerides Test. We also included the time to prediction (time between visits); gender; age at the repeated visit; and a bias term which is related to the prevalence in the population. We computed the values of these features’ associated coefficients with their 95%CI to enable a reconstruction of the models (Figure 3E).

As expected, the HbA1c% feature had the highest predictive power since it is one of the criteria for T2D diagnosis. The second-highest predictive feature was the high-light-scatter-reticulocytes-count, which reflects the number of new red blood cells in the body^23^. HDL cholesterol, which is known to be beneficial for health, especially in the context of cardiovascular diseases and T2D ^15,24,25^, was found to be inversely correlated to the predicted probability of T2D onset. Interestingly, age and sex had a very low OR value, meaning that they hardly contributed to the model, probably because the T2D relevant information of these features latent within the blood-tests’ data. Using the five blood tests scoreboard model, we removed the use of age, as we found that it did not contribute to the final score of the model.

### 2.3 Prediction within an HbA1c% stratified population

To verify that our models are capable of discriminating within a group of normoglycemic participants and within a group of pre-diabetic participants, we tested the models separately on each group extracted from our data. We separated the groups based on their HbA1c% levels during the first visit to the UKB assessment centers. We allocated participants with 4%<HbA1c%<=5.6% to the normoglycemic group, and participants with 5.7%<HbA1c%<6.5% levels to the pre-diabetic group^26^. As HbA1c% is one of the identifiers of T2D, this measure is a strong predictor of T2D. The prevalence of T2D onset within the normoglycemic group was only 1% versus a prevalence of 12% in the pre-diabetic group. We examined the driving factors of T2D in each of these stratified groups (Table 1S). Within the normoglycemic group, the anthropometry model yielded an auROC of 0.81 (0.76-0.85) with an APS of 0.05 (0.03-0.08) and deciles’ OR of x31 (8.2-51). When testing the models within the pre-diabetic group, the anthropometry model achieved an auROC of 0.75 (0.7-0.79), APS of 0.32 (0.24-0.41) and deciles’ OR of x26 (9.6-37). Both of these results outperform the FINDRISC and the GDRS results. For the normoglycemic HbA1c% range the anthropometry model resulted in an auROC of 0.82 (0.77-0.87), APS of 0.06 (0.04-0.1) and deciles’ OR of x29 (7.5-56). These results are similar to those of the five blood tests model’s results (Figure 3 A-C).

## 3. Discussion and conclusions

In this study, we analyzed several models for predicting the onset of T2D, which we trained and tested on a UKB based cohort, aged 40-69. Due to their accessibility and high predictability, we suggest two simple logistic regression models: the anthropometric and the five blood tests models. These models are suited for the UKB cohort, or populations with similar characteristics (See Table 1).

To provide an accessible and simple, yet predictive model, we based our first proposed model on eight non-laboratory anthropometric measures. We then developed an additional, straightforward model which is more accurate than the anthropometric model, to be used when laboratory blood tests are within reach. We based our second proposed model on five blood tests, including the age and sex of the participants. Both models are provided in their logistic regression form which is more accurate’ yet requires a computer aided analysis, and in an easy to use scoreboards form. For both models, we obtained results that were superior to those of the current clinically validated non-laboratory models, the Finnish Diabetes Risk SCore (FINDRISC) and the German Diabetes Risk Score (GDRS). To have a fair comparison, we trained these reference models and evaluated the predictions on the data sets of our models.

Our models achieved a better auROC, APS, and decile prevalence OR, and better-calibrated predictions than the FINDRISC and GDRS models. The anthropometrics model and the five blood tests logistic regression model delivered prevalence OR of x77 and x87, respectively, while their scoreboard forms achieved OR of x61 and x78 respectively.

Analysing our models’ feature importance, we conclude that the most predictive features of the anthropometry model are the waist to hip ratio (WHR) and body mass index (BMI), both of which are body measures that also encapsulate data regarding body type or shape. These features are known in the literature as being related to T2D, for instance in the metabolic syndrome^19^. The most predictive features of the five blood tests model are the HbA1c%, which is a measure of the glycated-haemoglobin carried by the red blood cells often used to diagnose Diabetes, and the Reticulocyte count which is a measure of the number of young red blood cells. Using both these features may provide a better indication of the average blood sugar level during the last two-three months than using just the standard HbA1C% measure. Interestingly, Age and Sex had a very low OR value, meaning that they hardly contributed to the model. One explanation might be that the T2D related information of these features is already latent within the blood test data. For instance, the SHBG feature contains a continuous measure regarding the sex hormone of each participant thus making the Sex feature redundant.

One of the limitations of our study is that our cohort is biased from the actual U.K. population. Our cohort’s T2D prevalence was only 2.16% during the time of the research, while the general UK population T2D prevalence is 6.3%, and 8% among adults aged 45-54 in the general UK population (2019)^27^. This bias is commonly reported as a “healthy volunteer” selection bias ^28,29^, which reduces the T2D prevalence from 6% in the general UK population to 4.8% in the UKB population. An additional screening bias is caused by including only healthy participants at the first visit. This reduced the prevalence of T2D in our cohort to 2.08%. As such, to apply our models to additional populations, further research on their ethnicity and fine tuning of the feature coefficients might be required.

As several studies have concluded ^7,8,9^, a healthy lifestyle and diet modifications before the inception of T2D are expected to reduce the probability of T2D onset. Therefore, identifying people at risk for this disease is crucial. We assert that our models make a significant contribution to such identification in two ways: The laboratory five blood tests model for clinical use is highly predictive of T2D onset, and the anthropometrics mode, mainly in its coreboard form, is an easily accessible and accurate tool. Thus these models carry the potential to improve millions of people’s lives and reduce the economic burden on the medical system.

## 4. Methods

### 4.1 Data

We analysed UKB’s observational data of 500,000 participants that were recruited voluntarily during 2006-2010 from across the UK, aged 40-69. During the baseline assessment visit to the UK Biobank, the participants self-completed questionnaires, including lifestyle and other potentially health-related information. The participants also went through physical and biological measurements. Out of this cohort, we used the data of 20,346 participants who revisited the UK Biobank assessment center during 2012-2013 during the longitudinal research, and we also used the data of 48,705 participants that revisited for a second or third visit from 2014 onwards for an imaging visit and went through a medical check very similar to the one in the first visit to the assessment centre. We performed a screening process on the participants to keep only the ones who were not treated nor have T2D. We thus kept data of 44,873 participants in our study cohort, from which 2.16% developed T2D during a follow-up period of 7.3±2.3 years (Table 1, Figure 1A).

We started with 798 features for each participant and removed all the features which had more than 50% missing data points in our cohort. We later removed from the cohort all the participants who still had more than 25% missing data points. We then imputed the remaining missing data. We further removed from the study the participants who self-reported as being healthy but had HbA1c% levels higher than the healthy level of glycated haemoglobin (HbA1c%) test, which is often used to identify T2D, measuring the average blood sugar for the past 2 to 3 months. As not all of the participants had HbA1c% measurements, we estimate the bias of participants reporting as being healthy while having an HbA1c% level indicating as being diabetic. To do so, we used the data we have from a subpopulation of our patients and found it to be 0.5% of participants who reported as being healthy with a median HbA1c% value of 6.7%, while the cutoff for having T2D is 6.5%. (Table 1)

### 4.2 Feature selection process

For the feature selection process, we started with 798 features that we estimated as potential predictors for T2D onset. We then removed all the features which had more than 50% missing data values, leaving 279 features for the research. Next, we imputed the missing data of the remaining records (See methods). As the genetic input for some of the models, we used for each participant both Polygenic Risk Scores (PRS) and Single-Nucleotide-Polymorphisms (SNPs) from the UKB SNP array (See methods). We used forty-one PRSs with 129±37.8 SNPs on average for each PRS. We also used the single SNPs of each PRS as some of the models’ features; after removal of duplicate SNPs, we remained with 2267 SNPs (See methods).

Out of the screened features and the genetic data, we aggregated the features into thirteen sperate groups: age and sex; genetics; early life factors; sociodemographics; mental health; blood pressure and heart rate; family and ethnicity; medication; diet; lifestyle and physical activity; physical health; anthropometry; blood tests. We then ran models for each group of features separately; later, we trained models where we added the features groups according to their marginal predictability. (Figure 1A, supplementary material).

After selecting our leading models from the training and validation data sets, we tested and reported the results of the selected models from the holdout test set (Figure S1, see methods, supplementary material). To encourage extensive clinical use of our models, we optimised the number of features we use. We chose the logistic regression models as our final models due to their simplicity and interpretability while providing similar results to the GBDT models that we validated (See methods). For the purpose of simplifying our models, we analysed the validation models’ features’ importance, and iteratively removed the least contributing features (See methods, supplementary material). We used each normalised feature’s coefficient as a measure of its importance in the model.

### 4.3 Outcome

Our models provide a prediction score for the participant risk of developing T2D during a specific timeframe. The mean time between the first visit and the prediction time point in our cohort is 7.3±2.3 years. The results that we report are of a holdout test-set comprising 20% of our cohort that we kept aside up until the final report of the results. We trained all the models using the same training set, and we then reported the test results of the holdout test-set. We present the area under the receiver operating curve (auROC) and also the average precision score (APS) as the main metrics of our models. Using these models, a physician can inform patients regarding the risk fold of developing T2D vs the participants in the lowest risk decile or vs any other risk decile. Using the scoreboards models, a patient can obtain its predicted risk of developing T2D during the following years.

We calibrate the models to enable reporting of the probability to develop a T2D during a given timeframe. Calibration refers to the concurrence between the real T2D onset occurrence in a subpopulation and predicted T2D onset probabilities in this population. Since our data is highly imbalanced, with the prevalence of 2.17% T2D, we used one thousand bootstrapping iterations of each model to better estimate the mean predicted value in each calibration bin. To calculate the calibration curves, we first split the prediction of each model to ten deciles bins in the range of zero to one. We then scale the results using SKlearn’s isotonic regression calibration with five-fold cross-validation ^30^. We do so for each of the bootstrapping iterations. We then concatenate all the calibrated results and calculate the overall mean predicted probability for each probability decile.

For the quantisation of the risk groups of the scoreboards model, we performed a similar bootstrapping process on our validation data set. We considered several potential risk score limits that separate T2D onset probability, and we chose boundaries that showed good separation between risk groups. We then measured the prevalence in each risk group on the test set and we report these results.

### 4.4 Missing data

After removing all features with more than 50% missing data, and removing all the data of participants with more than 25% missing features, we imputed the remaining missing data. We analysed the correlations between predictors with missing data and found mainly correlations within anthropometry group features to other features in the same domain and same for blood-tests. We used SKlearn’s iterative imputer with a maximum of 10 iterations for the imputation and a tolerance of 0.1^30^. We imputed the training and validation sets apart from the imputation of the holdout test-set. We did not perform imputation on the categorical features but rather transformed them into one hot encoding with a bin for missing data using Pandas categorical tools.

### 4.5 Genetic data

We use both Polygenic Risk scores (PRS) and Single-Nucleotide-Polymorphisms (SNPs) as genetic input for some of the models. We calculate the PRS by summing the top correlated risk allele effect-sizes derived from Genome-Wide Association Studies (GWAS) summary statistics. To do so we first extracted from each summary statistics the top 1000 SNPs according to their p-value. We then took only the SNP’s that were presented also in the UKB SNP-array. We used 41 PRSs with 129±37.8 SNPs on average for each PRS. We also used the single SNPs of each PRS as features for some of the models, after removal of duplicate SNPs, we kept 2267 SNPs as features. The full list of the PRS summary statistics is given in the supplementary material.

To prevent data leakage, we calculated the PRS scores according to summary statistics publicly available from studies that were not derived from the UK Biobank. We also provided the models which include genetic data and raw SNPs data as an input.

### 4.6 Baseline models

As the reference models for our results, we used the well established FINDRISC and GDRS models^14,17,18^, which we retrained and tested on the same data that we used for our models (See methods). These two models are based on the Finnish and German populations which are relatively close to the U.K. population and on similar age ranges.

We trained and tested these models on the same data that we use for our models. We derive a FINDRISC score for each participant using the data for age, sex, Body Mass Index (BMI), waist circumference, and blood pressure medication as provided from the UKB. To calculate the duration of the physical activity score, as required by the FINDRISC model, we summed up the values of “duration of moderate activity” and “duration of vigorous activity” as provided by the UKB. As a measure for the consumption of vegetables and fruits we summed up the categories “cooked vegetable intake”, “Salad/raw vegetable intake”, and “fresh fruit intake” categories from the UKB. As an answer regarding the question *“Have any members of your patient’s immediate family or other relatives been diagnosed with diabetes (type 1 or type 2)? This question applies to blood relatives only”* we used the fields for the illness of the mother, the father and the siblings of each participant.

We lacked the data regarding participant’s grandparents, aunts, uncles, first cousins and children. We also lack the data regarding past blood pressure medication, but rather have the data for the current medication usage. Following the calculation of the FINDRISC score for each participant, we trained a logistic regression model using the score for each participant as the model input, and the probability of developing T2D as the output. We also calculated an additional model, in which we added the time of the second visit as an input for the FINDRISC mode, but found no major differences between the two. We report here the results for the FINDRISC model without time of the second visit as a feature.

To derive the GDRS model, we built a Cox regression model using Python’s lifelines package ^32^. As the features for the GDRS model we calculated the following features: years between visits; height; prevalent hypertension; physical activity (h/week); smoking habits (Former smoker<20 units per day or >=20 units per day, current smoker >=20 units per day or <20 units per day); whole bread intake; coffee intake; red meat consumption; one parent with diabetes; both parents with diabetes and a sibling with diabetes. We performed a random hyperparameters search in the same way that we used for our models. The hyperparameters we used here are: the penaliser parameter in the range of 0-10 using a 0.1 resolution; variance threshold 0-1 with 0.01 resolution to drop columns where the variance of the column was lower than the variance threshold.

### 4.7 Model building procedures

To prevent overfitting and biased models, we split the data to twenty percent of a holdout test set which we used only for the final reporting of results. From the remaining data, we split again into a thirty percent validation set and a seventy percent for the training set. We then use a two-stage process to evaluate the models’ performance: an exploration phase and a test phase (Figure 1, S1). During the exploration stage, we select the optimal features for our models using the training and validated data sets. For each group of features, we optimised the hyperparameters using two-hundred iterations of a random selection process. In each iteration, we measured the performance using the auROC metric with a five-fold cross-validation within the training set.

We later trained a model on the full training set with the top ranked hyper-parameters from the previous step. We test this model using the validation data set. We use this stage to compare various models and for the features selection process for our models.

At the final phase, the test phase, we report the results of our selected models. In this phase, we evaluate the selected models on the holdout test-set. To do so, we rerun the hyperparameters selection process using the training and validation data sets. We train the selected models with the selected hyperparameters on the pooled training and validation data sets. Lastly, we calculate the results of the trained model based on the holdout test-set. We use the same datasets for all of the discussed models.

For the logistic regression models we used SKlearn’s LogisticRegressionCV model ^30^. For the GBDT models we used Microsoft’s LightGBM package ^33^, and for the survival analysis models, we used the lifelines package ^32^.

During the models’ calculation process we used two-hundred iterations of random hyperparameters-search for the training of the models. For the GBDT models we used the following parameters values for the search: number of leaves - [2, 4, 8, 16, 32, 64, 128]; Number of boosting iterations - [50, 100, 250, 500, 1000, 2000, 4000]; learning rate - [0.005, 0.01, 0.05]; minimum child samples - [5, 10, 25, 50]; subsample - [0.5, 0.7, 0.9, 1]; features fraction - [0.01, 0.05, 0.1, 0.25, 0.5, 0.7, 1]; lambda l1 - [0, 0.25, 0.5, 0.9, 0.99, 0.999]; lambda l2 - [0, 0.25, 0.5, 0.9, 0.99, 0.999]; bagging frequency - [0, 1, 5]; bagging fraction- [0.5, 0.75, 1] ^33^.

For the logistic regression models, during the hyperparameters search we used penaliser at the raNGE OF 0-2 with 0.02 resolution for the l2 penalty.

### 4.8 SHAP

As the feature importance analysis for the GBDT model, we used the SHAP method, which approximates Shapley values. SHAP (SHapley Additive exPlanations) originated in a game theory, intended to explain the output of any machine learning model. SHAP Approximates the average marginal contributions of each feature of a model across all permutations of the other features in the same model ^34^.

### 4.9 Predictors

To estimate the contribution of each feature’s domain and for initial screening of features, we started by building a GBDT model based on 279 features plus genetics data originating from the UKB SNPs array. We used T2D related summary statistics from Genome-Wide-Association-Studies (GWAS). These are genetic studies designed to find correlations between known genetic variants and a phenotype of interest. To avoid data leakage, we used only GWASs that derived from outside the UKB population (See supplementary material for the full list of PRSs). As the feature importance analysis for the GBDT model, we used the SHAP method ^34^, which approximates Shapley values (See methods).

To select the most predictive features for the anthropometry and the blood-tests models, we trained and tested the full-features model using the training and validation cohort, and then used this model’s feature importance to extract the most predictive features. We also analysed models which included data of family relatives with T2D using only the training and validation sets. As we did not observe any major improvement over the anthropometrics model, for the simplicity of the model, we decided to omit this feature. At the last step, we tested and reported the model on the holdout test.

For the extraction of the five blood tests model, we performed a features selection process by evaluating logistic regression models using the training and validation datasets. We ran models with twenty, ten, and down to four features of blood tests together with age and sex as features, each time removing the blood test with the least essential feature score. We then selected the model with five blood tests (HbA1c%, Reticulocytes count, Gamma Glutamyl Transferase (GGT), Triglycerides, HDL cholesterol, age and sex) as the optimal balance between model’s simplicity (low number of features) and model’s accuracy (using more features) and report its results on the holdout test set.

We normalised all the continuous predictors using the standard z-score. In order to avoid data leakage, the train-validation sets were normalised apart from the holdout test set.

### 4.10 Models calibration

For each of the models, we calculated the deviation of the mean predicted probability from the actual T2D prevalence of each bin.

We split the probabilities range (0-1) to ten prediction probabilities bins with probabilities resolution of 0.1 (Figure 3F). We assign each prediction’s sample to a decile bin according to the calibrated predicted probability of T2D onset. Since our data is highly imbalanced, with a prevalence of 2.17% T2D onset, we used one thousand bootstrapping iterations to better calibrate the models. As such, each participant might be present at several bins according to each prediction iteration of the bootstrapping process. We repeated this process also for the scoreboards models.

### 4.11 Extracting scoreboards

To extract our scoreboards, we explored the train and validation data sets, and reported the results on the hold out data set. We calculated the weight of evidence (WoE) of our data by splitting each of our features into bins. We binned in higher resolution features that have greater importance, while maintaining monotonically increasing WoE. (Anon n.d.) For the quantisation of the risk groups of the scoreboards model, we performed one thousand iterations of the bootstrapping process on our validation data set. We considered several potential risk score limits that separate T2D onset probability in each of the scores groups, and we chose boundaries that showed a separation between the risk groups. We then measured the prevalence in each risk group on the test set and we report these results.

### 4.12 References for PRS summary statistics articles

HbA1c^353,36,37^; Cigarettes per day, ever smoked, age start smoking^38^; HOMA-IR, HOMA-B, diabetes BMI unadjusted, diabetes BMI adjusted, fasting glucose ^39^; Fasting glucose, 2 hours glucose level,fasting insulin, fasting insulin adjusted BMI’-(MAGIC_Scott)^40^; Fasting glucose, fasting glucose adjusted for BMI,fasting insulin adjusted for BMI^41^; Two hours glucose level^42^; Fasting insulin ^43^; Fasting Proinsulin^44^; Leptin adjusted for BMI, Leptin unadjusted for BMI^45^; Triglycerides, Cholesterol, ldl, hdl^46^; BMI^47^; Obesity class1, obesity_class2, overweight ^48^;Anorexia^49^; Height^50^; Waist circumference, hips circumference^51^; Cardio^52^; Heart_Rate^53^; Alzheimer^54^; Asthma ^55^

## Data Availability

The UKB data are available through the UK Biobank Access Management System https://www.ukbiobank.ac.uk/

## 5. Acknowledgements

This research has been conducted using the UK Biobank Resource under Application Number 28784

