## Supplementary material for "Prediction of type 2 diabetes mellitus onset using logistic regression-based scoreboards"

#### Figure S1 - Models testing and training process

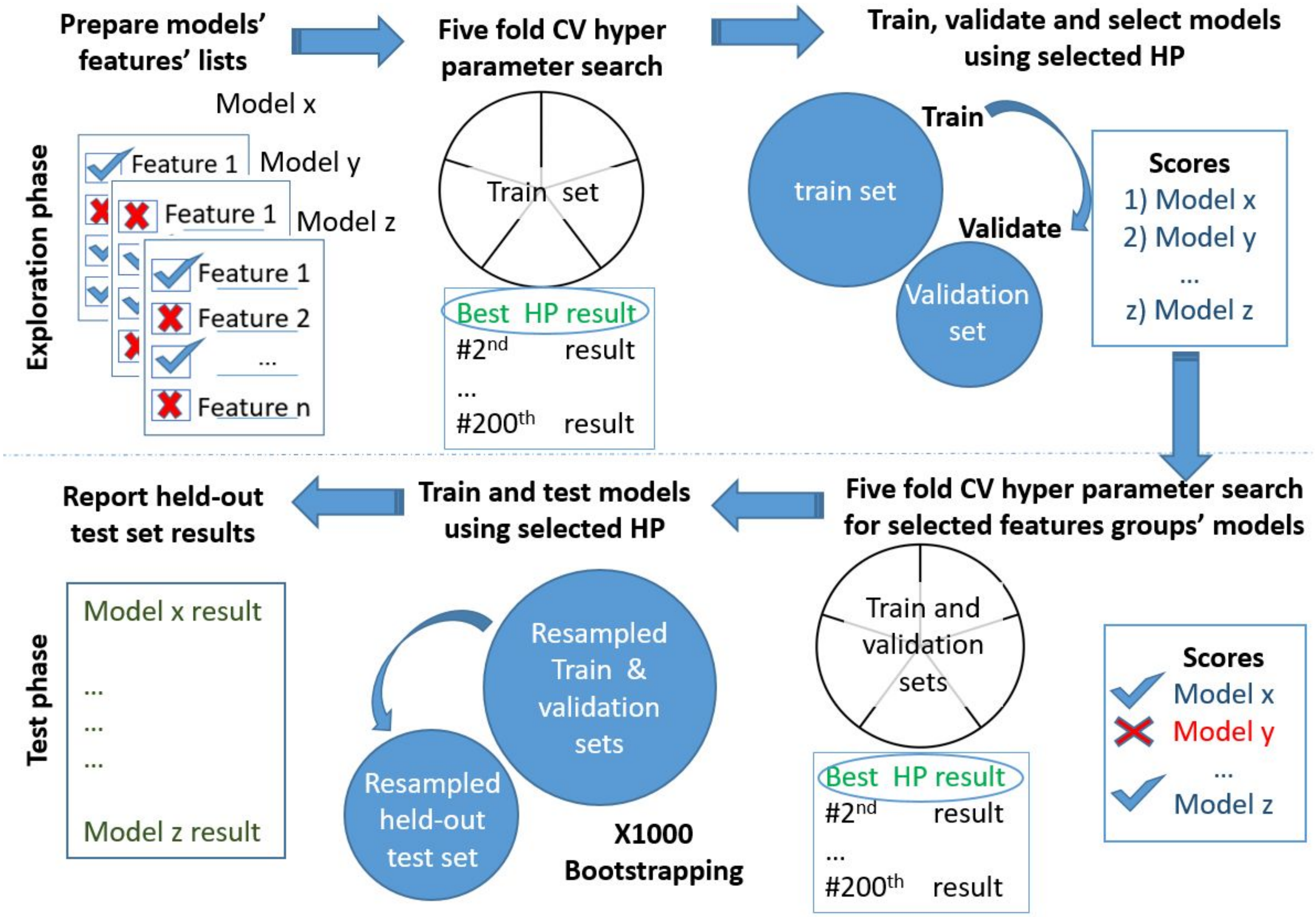


**Figure S1 |Models' extraction flow**. Scheme of the models' exploration and evaluation process. We use a fivefold CV with two hundred iterations of random hyperparameters (HP) selection process for each of the features groups. We then select the top-scored HP for each feature's group and Train a new model based on the train set and measure the auROC using the validation set. Out of the models validated, we chose the models that comply with objectives of minimal features and high performance. The reported results are of the held-out test set.

### Prediction within an HbA1c% stratified population

To verify that our model not only discriminates between participants who were already having elevated HbA1c% levels but is also capable of discriminating within a group of participants with normal or prediabetes HbA1c% levels we stratified our data to normoglycemic and prediabetic groups according to their HbA1c% levels. As patients are diagnosed by their HbA1c% levels, this measure is a strong predictor of T2D in itself with a 12% prevalence within the group of participants with HbA1c% levels 5.7-6.5%, versus only 1% of T2D onset for participants with HbA1c% levels 4-5.7% in our cohort.

We tested our models on these two T2D risk populations from our cohort. We examine what are the driving factors of T2D in each of these stratified groups (Figure S2). Within the normoglycemic group, the anthropometry model provides auROC of 0.81 (0.76-0.85) with an APS of 0.05 (0.03-0.08) and deciles' OR of x31 (8.2-51). These results outperform the FINDRISC results with auROC, APS and deciles' OR of 0.7 (0.65-0.76), 0.03 (0.02-0.04), and x20 (5.2-42) respectively. These results also surpass the GDRS results with auROC, APS, and deciles' OR of 0.61 (0.55-0.67), 0.02 (0.01-0.02), and x14(3.3-25) respectively. The Anthropometry model results in this normal HbA1c% range are similar to those of the five blood tests model results with auROC of 0.82 (0.77-0.87), APS of 0.06 (0.04-0.1) and deciles' OR of x29 (7.5-56). (Figure 2S A-C).

When we test the models within the pre-diabetic group, the anthropometry model achieves auROC of 0.75 (0.7-0.79), APS of 0.32 (0.24-0.41) deciles' OR of x26 (9.6-37). These results are again superior to the FINDRISC results with auROC, APS, and deciles' OR of 0.66 (0.61-0.71), 0.21 (0.16-0.26), and x7.4(2.5-21) respectively. It also surpasses the GDRS results with auROC, APS and deciles' OR of 0.58 (0.53-0.63), 0.16 (0.12-0.2) ,and x1.9 (0.61-5.5) respectively. In this HbA1c% range, the five blood tests model outperform the Anthropometry model with auROC, APS, and deciles' OR of 0.84 (0.80-0.87), 0.48 (0.38-0.57), and x38 (29-47) respectively. (Figure 2S a-c).

Comparing the feature importance of the Anthropometry model in the normoglycemic and pre-diabetic subpopulations indicates that the main differences are in the bias coefficient, which reflects the higher prevalence of T2D in the pre-diabetic group and the time between visits feature is also becoming more dominant than waist to hips ratio and BMI. Suggesting that in the pre-diabetic group, it is mostly a matter of time until T2D emerges. Within the normoglycemic group, we see that the most predictable feature is the WHR, followed by BMI - both taking into account not only the body measures but also implicit body measures ratios which indicate about the effect of the body shape (Figure S2 D). When we compare the five-blood-tests model's features importance, it appears that the features importance values did not vary much apart from the negative impact of HDL cholesterol in the normoglycemic group and the bias term which indicates the higher prevalence in the pre-diabetic group (Figure S2-E).

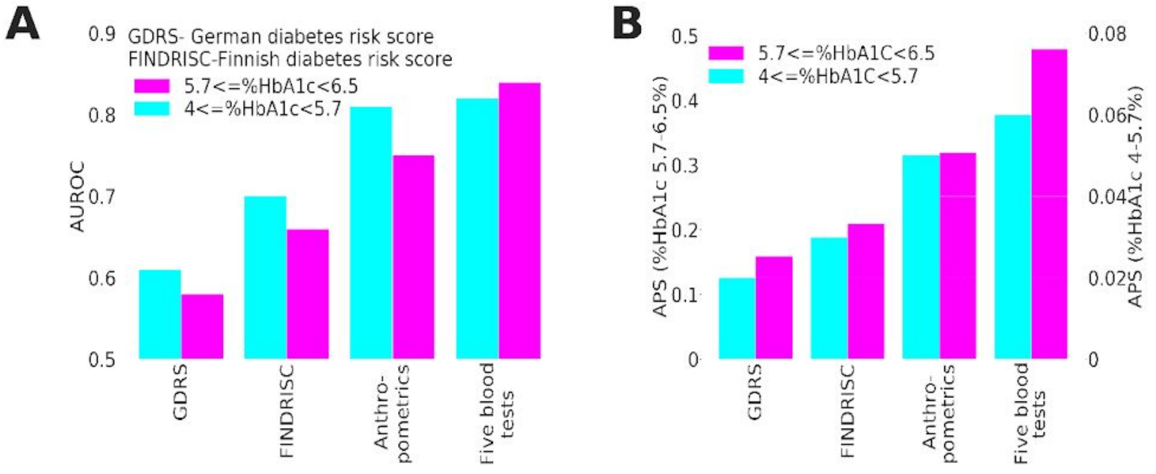


**C**

| | Train set | Test set | Label | APS | AUROC | Deciles fold |
|---|---|---|---|---|---|---|
| **HbA1c% 5.7%-6.5%** | n=3504 Prevalence= 12.7% | n=914 Prevalence= 12.8% | Five Blood tests | 0.48 (0.38-0.57) | 0.84 (0.80-0.87) | 38 (29-47) |
| | | | Anthropometry | 0.32 (0.24-0.41) | 0.75 (0.70-0.79) | 26 (9.6-37) |
| | | | FINDRISC | 0.21 (0.16-0.26) | 0.66 (0.61-0.71) | 7.4 (2.5-21) |
| | | | German diabetes risk score GDRS | 0.16 (0.12-0.20) | 0.58 (0.53-0.63) | 1.9 (0.61-5.5) |
| **HbA1c% 4-5.7%** | n=32,352 Prevalence= 1% | n=8078 Prevalence= 1% | Five Blood tests | 0.06 (0.04-0.10) | 0.82 (0.77-0.87) | 29 (7.5-56) |
| | | | Anthropometry | 0.05 (0.03-0.08) | 0.81 (0.76-0.85) | 31 (8.2-51) |
| | | | FINDRISC | 0.03 (0.02-0.04) | 0.70 (0.65-0.76) | 20 (5.2-42) |
| | | | German diabetes risk score | 0.02 (0.01-0.02) | 0.61 (0.55-0.67) | 14 (3.3-25) |

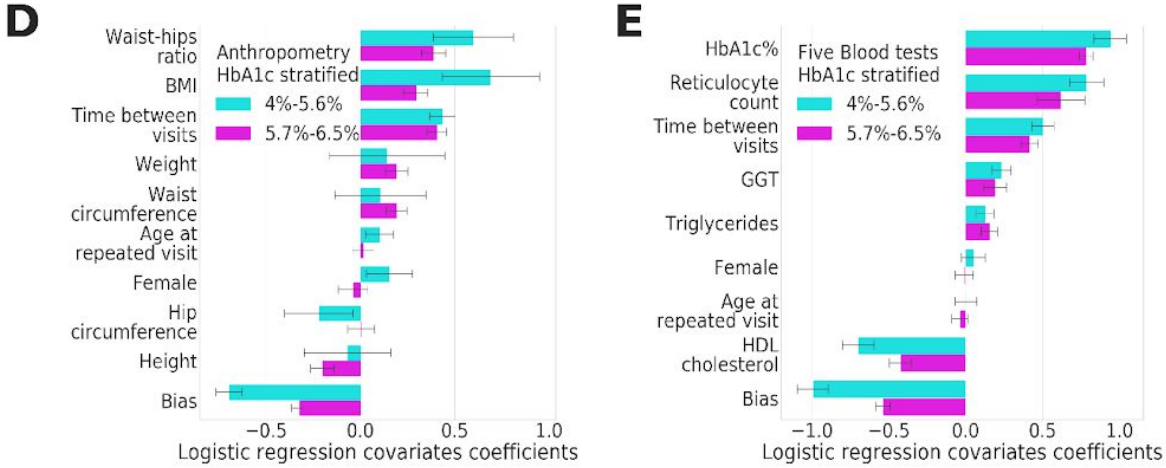


**Figure S2 | Stratified populations according to HbA1c% - models comparison**. **A)** auROC in both normoglycemic and pre-diabetic groups. Although the five-blood-tests model achieves more accurate scores than the anthropometric model in both populations, the anthropometry model's results approach the five blood tests model in the normoglycemic group. Both models outperform the FINDRISC and GDRS models.

**B) Comparison of model's APS within the HbA1c% stratified groups.** Left Y-axis indicates the APS of the models' in the prediabetics group, the right Y-axis indicates APS in the normoglycemic group, we see again that the order of model's predictability remains Five blood tests; Anthropometry; FINDRISC and lastly the GDRS model. **C)** A summary table for the various models AuROC, Average-Precision and deciles risk fold scores for the HbA1c% stratified subpopulations. **D)** A feature importance comparison of the Anthropometric model within the HBbA1c% stratified groups. showing a stronger impact of WHR and BMI in the normoglycemic group and a stronger negative impact of the bias - indicating the higher prevalence in the pre-diabetic group. **E)** A feature importance comparison of the five-blood-tests model within the HBbA1c% stratified groups. showing a stronger negative impact of HDL cholesterol in the normoglycemic group and a stronger negative impact of the bias - indicating the lower prevalence in the normoglycemic group.

Comparing the five blood tests model's features importance, the features importance values do not vary much. The main differences are the stronger negative impact of HDL cholesterol on the prediction results and the trivial bias term, which indicates the lower prevalence in the normoglycemic group (Figure S2 E).

#### Exploring the full features' space using gradient boosted decision trees.

To select features for our simple models, we analysed the features' importance of features that we sought of having some relation to T2D. We analyse what is the power of a predictive model with a vast amount of information, to compare it to our minimal features models.

We started by sorting out a list of 279 covariates from the first visit to the UKB assessment centre to be used as our preliminary features. On top of these features, we used the UKB single-nucleotide polymorphisms (SNPs) genotyping data and their calculated PRS.

We inspected the impact of various features' groups, using the lightGBM [30] gradient decision trees model using SHAP [32,31] (See methods). We aggregated the features into thirteen seperate groups: age and sex; genetics; early life factors; sociodemographics; mental health; blood pressure and heart rate; family and ethnicity; medication; diet; lifestyle and physical activity; physical health; anthropometry; blood tests. All of these groups included age and sex.

We also tested the impact of HbA1c% with sge and dex; genetics without age and Sex (Table S1). The top five predictive GBDT models, according to their auROC, APS, and decile folds, in descending order are: the "All features" model with auROC 0.9 (0.88-0.92 95%CI), APS of 0.28 (0.22-0.35 95% CI) and a deciles' OR of x65 (49-73); The "full-blood-tests" model with auROC 0.89 (0.86-0.9), APS 0.25 (0.2-0.31), and deciles' OR x62 (48-69); The "five blood tests" model with auROC 0.87(0.85-0.89), APS 0.21 (0.17-0.27) and deciles' OR of x59(49-66); the HbA1c% based model with auROC 0.81 (0.78-0.84), APS 0.18 (0.13-0.23) and deciles' OR of x32(11-59); and the anthropometry model with auROC 0.76 (0.73-0.79), APS 0.07 (0.06-0.1) and deciles' OR x30 (12-42) (Table 1S).

To explore the heritability impact on the predictability of the models, we used two main features groups. One is a broad sense heritability based on SNPs and PRSs, and the other one is heritability which includes family and ethnicity. Adding the SNPs and PRSs data to the age and sex group, elevated the APS and auROC from 0.03 (0.02-0.04), 0.58 (0.54-0.62) to 0.04 (0.03-0.05) and 0.61 (0.57-0.65) respectively.

Adding this DNA array data to all other features together did not provide a considerable contribution to prediction. Using the family and ethnicity features with age and sex provides an APS of 0.04 (0.03-0.05), auROC of 0.63(0.58-0.67), and deciles' OR of x5.4 (2-14) (Table S1), considerably lower than other features' groups such as the anthropometry group with auROC 0.76(0.73-0.79) and deciles' OR of x30 (12-42) and from the lifestyle and physical activity group, scoring auROC of 0.69 (0.65-0.72) and deciles' OR of x16 (6-29). A possible explanation for the low predictability of heritable features is that T2D genetics plays a role mainly in the younger population and the greater role played by environmental factors in those who develop diabetes late in life [33]

Another interesting result is a lifestyle and physical activity model, which includes ninety-eight features related to: physical activity; addictions; alcohol, smoking and cannabis use; electronic device use; employment; sexual factors; sleeping; social support and sun exposure. This model achieves an auROC of 0.69 (0.65-0.72) and deciles' OR of x16 (6-29), and it provides better prediction scores than the diet features' group. This model includes thirty-two diet features from the UKB touchscreen questionnaire on the reported frequency of type and intake of a range of common food and drink items. The diet based model achieved auROC of 0.64 (0.6-0.67) and deciles' OR of x6 (2-17) (Table S1).

| Label | APS | AUROC | Deciles fold |
|---|---|---|---|
| **All** | 0.28 (0.22-0.35) | 0.90 (0.88-0.92) | 65.48 (49-73) |
| **Blood Tests** | 0.25 (0.20-0.31) | 0.89 (0.86-0.90) | 62.18 (48-69) |
| **Five blood tests** | 0.21 (0.17-0.27) | 0.87 (0.85-0.89) | 59.30 (49-66) |
| **HbA1c%** | 0.18 (0.13-0.23) | 0.81 (0.78-0.84) | 31.60 (11-59) |
| **Anthropometry** | 0.07 (0.06-0.10) | 0.76 (0.73-0.79) | 29.84 (12-42) |
| **Physical health** | 0.05 (0.04-0.07) | 0.69 (0.65-0.73) | 15.44 (5.-29) |
| **Lifestyle and physical activity** | 0.05 (0.04-0.07) | 0.69 (0.65-0.72) | 16.40 (6.-29) |
| **Diet** | 0.04 (0.03-0.05) | 0.64 (0.60-0.67) | 6.03 (2.-17) |
| **Medication** | 0.04 (0.03-0.05) | 0.63 (0.59-0.67) | 5.60 (2.-14) |
| **Family and Ethnicity** | 0.04 (0.03-0.05) | 0.63 (0.58-0.67) | 5.35 (2.-14) |
| **BP and HR** | 0.04 (0.03-0.05) | 0.62 (0.58-0.66) | 8.54 (3.-20) |
| **Mental health** | 0.04 (0.03-0.05) | 0.62 (0.57-0.66) | 6.12 (2.-17) |
| **Socio demographics** | 0.04 (0.03-0.05) | 0.61 (0.57-0.65) | 5.19 (2.-15) |
| **Genetics Age and Sex** | 0.04 (0.03-0.05) | 0.61 (0.57-0.65) | 5.31(2.-13) |
| **Early Life Factors** | 0.03 (0.02-0.04) | 0.58 (0.54-0.62) | 2.57 (1.1-5.9) |
| **Age and Sex** | 0.03 (0.02-0.04) | 0.58 (0.54-0.62) | 2.16 (0.9-4.4) |
| **Only genetics** | 0.03 (0.02-0.04) | 0.56 (0.52-0.60) | 2.50 (1.1-5.5) |

*Table S1:Predicting using features domain groups, Results table of GBDT models for various features domains. The logistic regression models provided better results than the GBDT models for the blood tests and anthropometrics based models.*

We then examined the additive contribution of each predictive group to the total predictive power of the "all features" model (Figure S4-A). We started with the baseline model of "age and sex" and added models one

after the other, sorted by their predictive power as a single features' group using GBDT. We conclude that using the five-blood-test model substantially increase the prediction results when compared to a model based only on HbA1c% with age, and sex.
The auROC, APS and deciles' OR increases from 0.81(0.78-0.84), 0.18(0.13-0.23) and x32(11-59) to 0.87(0.85-0.89), 0.21(0.17-0.27) and x59(49-66) respectively.

When we use the full blood tests model, the performance slightly increases to auROC, APS, and deciles' OR of 0.89(0.86-0.9), 0.25(0.2-0.31), and x62(48-69) respectively. We did not identify any major increase in results adding any other specific group to this list, suggesting that most of the predictive power of our models are captured by the blood test features or has collinearity with it. Using all features together provided an increase of performance to auROC, APS, and the deciles' OR of 0.9(0.88-0.92), 0.28(0.22-0.35), and x65(49-73) respectively (Figure S4 A-B).

To get some insights for the features that drive the T2D prediction, we built an additional model using all features excluding the HbA1c% and non-fasting glucose, as these features are highly predictive and might screen out other clinically interesting covariates. The auROC, APS, and the deciles' prevalence OR results that we achieved using this model are 0.13 (0.1-0.16), 0.84 (0.82-0.86), and x49(41-56), respectively.

To gain understanding into the features that contribute the most to the GBDT model predictions, we used the SHAP package feature importance framework. SHAP uses Shapley values, which are essentially the average marginal contribution of a feature over all possible combinations (see methods)[31] .

The top ten important features include Gamma-glutamyl-transferase (GGT) (1st in the list), and Alanine-aminotransferase (ALT) (9th in the list), positively related to T2D onset prediction; high levels of these enzymes usually signal for liver damage, which might occur from fatty liver. Increased values of waist circumference (2nd), body mass index (BMI) (3rd), and weight (10th) are all in the ten leading predictors for T2D in this model. Sex hormone-binding globulin (SHBG) (4th), which is a protein produced by the liver and transports sex hormones blood, is also related to a decreased probability of developing T2D in our model. This result aligns with research showing that raised SHBG levels reduce the risk of T2D [34]. High levels of triglycerides (5th), which is long known to precede T2D [35] , also drive the prediction of developing T2D in this model. High levels of HDL (8th) cholesterol reduce the probability of developing T2D in this model, which aligns with the literature findings regarding correlations; although, a genetic mendelian randomization study found it as correlated but non-causal for T2D [36,37]

Having no vascular or heart problems diagnosed reduces the probability of developing T2D. According to the National Health Interview Survey (NHIS) published in 1989, 14% of the diabetic population at the ages 45-64, also reported having ischemic heart disease, and both conditions are known to be related to the metabolic syndrome [38,39,40] . The time between visits, i.e., time from taking the features measurements to time of prediction is also in the top ten list, which implies that the cumulative probability of developing T2D increases over time.

Two features that are absent from this list are age and sex. While SHBG might confound sex - age seems to be irrelevant once we provide a model with the blood test results as features, and probably reflect a T2D "biological age" of the participant [41,42](Figure S4-C)

A

| Label | APS | AUROC | Deciles fold |
|---|---|---|---|
| 1)Age and Sex | 0.03 (0.02-0.04) | 0.58 (0.54-0.62) | 2.16 (0.94-4.3) |
| 2) +HbA1c% | 0.18 (0.13-0.23) | 0.81 (0.78-0.84) | 31.60 (11-59) |
| 3)Five blood tests | 0.21 (0.17-0.27) | 0.87 (0.85-0.89) | 59.30 (49-66) |
| 4) Blood Tests | 0.25 (0.20-0.31) | 0.89 (0.86-0.9) | 62.18 (48-69) |
| 5) + Anthropometric | 0.24 (0.19-0.3) | 0.89 (0.87-0.91) | 62.80 (54-70) |
| 6) + Physical Health | 0.27 (0.21-0.33) | 0.9 (0.88-0.92) | 64.37 (57-71) |
| 7) + Lifestyle | 0.27 (0.21-0.34) | 0.9 (0.88-0.92) | 65.34 (58-72) |
| 8) + Blood pressure and heart rate | 0.26 (0.19-0.32) | 0.89 (0.87-0.91) | 64.15 (57-71) |
| 9) + medical diagnosis (Non-diabetic) | 0.25 (0.20-0.32) | 0.9 (0.88-0.92) | 64.71 (57-72) |
| 10) + Mental health | 0.24 (0.18-0.3) | 0.89 (0.87-0.91) | 61.67 (48-69) |
| 11) + medication | 0.25 (0.20-0.31) | 0.9 (0.88-0.92) | 64.43 (58-71) |
| 12) + Diet | 0.27 (0.21-0.33) | 0.89 (0.87-0.91) | 64.37 (57-71) |
| 13) + Family and ethnicity | 0.25 (0.19-0.32) | 0.89 (0.87-0.91) | 63.66 (53-71) |
| 14) + Early life factors | 0.28 (0.22-0.34) | 0.9 (0.87-0.92) | 64.76 (58-72) |
| 15) + Sociodemographic | 0.26 (0.21-0.33) | 0.9 (0.88-0.92) | 65.11 (58-72) |
| 16) + Genetic sequencing | 0.28 (0.22-0.35) | 0.9 (0.88-0.92) | 65.48 (49-73) |

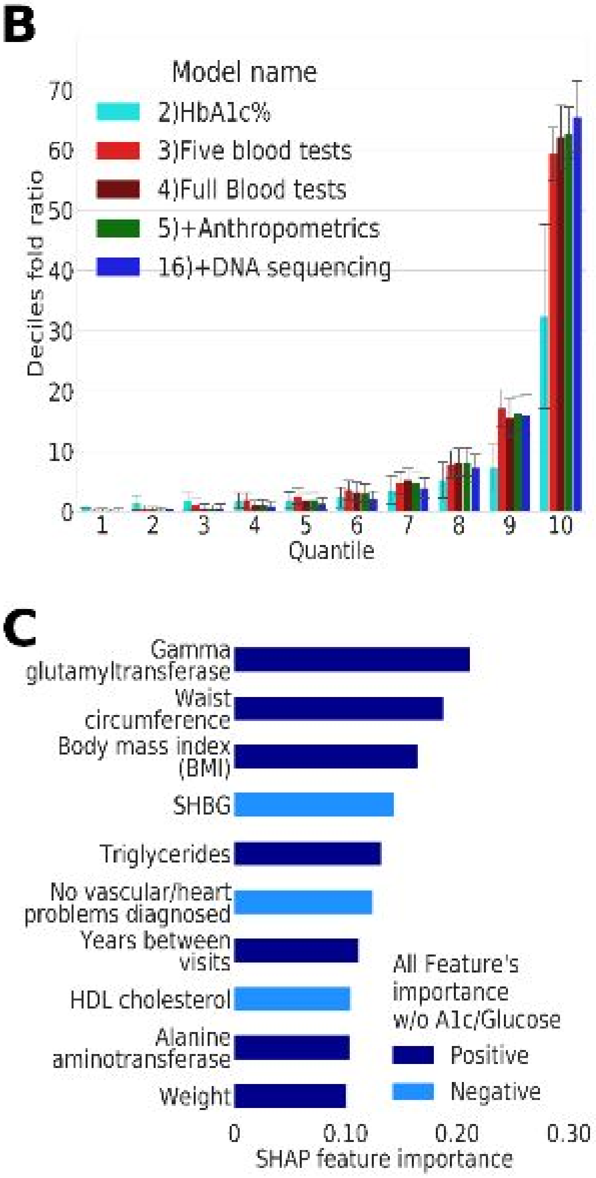


**Figure S4 | Summary of Incremental feature's model**: **A**. Comparison table of APS, auROC, and deciles' OR metrics for GBDT models, where each model is embedding the preceding model's features plus additional features domain. The largest increase in prediction is upon adding the HbA1C% feature, which is also a biomarker of T2D diagnosis. Adding the DNA sequencing data did not contribute much to the prediction power of the model. **B.** Deciles-prevalence-fold-ratio (DFPR) graph . The five blood tests model considerably outperforms the HbA1C% model. **C.** Shapley values feature's importance of the "All features without HbA1c% nor glucose" GBDT model. Removal of the HbA1c% and non-fasting glucose enables other major contributors to T2D prediction to be seen having an impact on T2D prediction.

### References for PRS summary statistics articles.

HbA1c[43,44,45] ; Cigarettes per day, ever smoked, age start smoking[46] ; HOMA-IR, HOMA-B, diabetes BMI unadjusted, diabetes BMI adjusted, fasting glucose [47]; Fasting glucose, 2 hours glucose level,fasting insulin, fasting insulin adjusted BMI'-(MAGIC_Scott)[48]; Fasting glucose, fasting glucose adjusted for BMI,fasting insulin adjusted for BMI[49]; Two hours glucose level[50]; Fasting insulin [51]; Fasting Proinsulin[52]; Leptin adjusted for BMI, Leptin unadjusted for BMI[53]; Triglycerides, Cholesterol, ldl, hdl[54]; BMI[55]; Obesity class1, obesity_class2, overweight [56];Anorexia[57]; Height[58]; Waist circumference, hips circumference[59]; Cardio[60]; Heart_Rate[61]; Alzheimer[62]; Asthma [63]

### Acknowledgements:

This research has been conducted using the UK Biobank Resource under Application Number 28784
